# Characterizing the effective reproduction number during the COVID-19 epidemic: Insights from Qatar’s experience

**DOI:** 10.1101/2021.10.07.21264599

**Authors:** Raghid Bsat, Hiam Chemaitelly, Peter Coyle, Patrick Tang, Mohammad R. Hasan, Zaina Al Kanaani, Einas Al Kuwari, Adeel A. Butt, Andrew Jeremijenko, Anvar Hassan Kaleeckal, Ali Nizar Latif, Riyazuddin Mohammad Shaik, Gheyath K. Nasrallah, Fatiha M. Benslimane, Hebah A. Al Khatib, Hadi M. Yassine, Mohamed G. Al Kuwari, Hamad Eid Al Romaihi, Mohamed H. Al-Thani, Abdullatif Al Khal, Roberto Bertollini, Laith J. Abu-Raddad, Houssein H. Ayoub

**Author notes:** **Reprints or correspondence:** Dr. Houssein H. Ayoub, Department of Mathematics, Statistics, and Physics, Qatar University, P.O. Box 2713, Doha, Qatar., Professor Laith J. Abu-Raddad, Infectious Disease Epidemiology Group, World Health Organization Collaborating Centre for Disease Epidemiology Analytics on HIV/AIDS, Sexually Transmitted Infections, and Viral Hepatitis, Weill Cornell Medicine - Qatar, Qatar Foundation - Education City, P. O. Box 24144, Doha, Qatar. Joint senior authors (Laith J. Abu-Raddad, Houssein H. Ayoub).

## Abstract

**Background:** The effective reproduction number, *R*_*t*_, is a tool to track and understand epidemic dynamics. This investigation of *R*_*t*_ estimations was conducted to guide the national COVID-19 response in Qatar, from the onset of the epidemic until August 18, 2021.

**Methods:** Real-time “empirical” 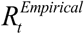 was estimated using five methods, including the Robert Koch Institute, Cislaghi, Systrom-Bettencourt and Ribeiro, Wallinga and Teunis, and Cori et al. methods. *R*_t_ was also estimated using a transmission dynamics model 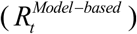. Uncertainty and sensitivity analyses were conducted. Agreements between different *R*_*t*_ estimates were assessed by calculating correlation coefficients.

**Results:** 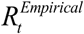 captured the evolution of the epidemic through three waves, public health response landmarks, effects of major social events, transient fluctuations coinciding with significant clusters of infection, and introduction and expansion of the B.1.1.7 variant. The various estimation methods produced consistent and overall comparable 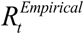 estimates with generally large correlation coefficients. The Wallinga and Teunis method was the fastest at detecting changes in epidemic dynamics. 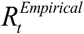 estimates were consistent whether using time series of symptomatic PCR-confirmed cases, all PCR-confirmed cases, acute-care hospital admissions, or ICU-care hospital admissions, to proxy trends in true infection incidence. 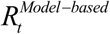 correlated strongly with 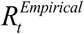 and provided an average 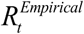.

**Conclusions:** *R*_*t*_ estimations were robust and generated consistent results regardless of the data source or the method of estimation. Findings affirmed an influential role for *R*_*t*_ estimations in guiding national responses to the COVID-19 pandemic, even in resource-limited settings.

## INTRODUCTION

The severe acute respiratory syndrome coronavirus 2 (SARS-CoV-2) pandemic is the most serious global health challenge in recent history [1,2]. Coronavirus Disease 2019 (COVID-19) morbidity and mortality has imposed unparalleled burdens on healthcare systems worldwide, and necessitated unprecedented restrictions on mobility and on social and economic activities [3,4]. Tracking and monitoring each wave of infection have become essential to avoid the adverse consequences of infection transmission [5-8]. With such serious consequences to the healthcare system, economy, and society, decisions regarding the escalation or easing of restrictions have become a critical facet of policymaking since the discovery of the virus in December of 2019 [6,9,10].

The effective reproduction number (*R*_*t*_), the average number of secondary infections each infection is generating at a given point in time [6,11-13], has proven to be one of the most influential tools in monitoring and tracking the epidemic, and informing the escalation and easing of restrictions [6,11-13]. In the present study, we report on the use of *R*_*t*_ in Qatar, to understand epidemic dynamics and to establish national policy decisions and public heath interventions, in what has become a successful country experience.

Qatar is a peninsula in the Arabian Gulf with a diverse population of 2.8 million people [5,14] that has been affected by three SARS-CoV-2 epidemic waves [5,6,15-19]. The first wave started with the introduction of the virus in February of 2020 and peaked in late May 2020 [5,6]. The second wave started in mid-January, 2021, and was triggered by the introduction and expansion of the B.1.1.7 (Alpha [20]) variant [15-19,21]. This wave peaked in the first week of March, but was followed immediately by a third wave that was triggered by introduction and rapid expansion of the B.1.351 (Beta [20]) variant, which started in mid-March and peaked in mid-April, 2021 [15-19,21].

Two forms of *R*_*t*_ estimation have been used in Qatar to inform the national COVID-19 response, and each proved to have its own intrinsic public health value. The first is the real-time “empirical” estimation which is done by calculating *R*_*t*_ directly from diagnosed cases. Different methods were explored for estimating the empirical *R*_*t*_ (henceforth, 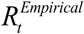), and based on this exploration the Robert Koch Institute method [13,22] was used for feasibility, ease of use, and functionality. The second estimation method was model-based by calculating *R*_*t*_ using a population-level compartmental transmission dynamics model [6,23], hereafter designated as 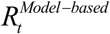.

## METHODS

### Data sources

Analyses were conducted using the centralized, integrated, and standardized national SARS-CoV-2 databases compiled at Hamad Medical Corporation (HMC), the main public healthcare provider and the nationally designated provider for all COVID-19 healthcare needs. These databases have captured all SARS-CoV-2-related data since the start of the epidemic, including all records of polymerase chain reaction (PCR) testing, antibody testing, COVID-19 hospitalizations, vaccinations, infection severity classification per World Health Organization (WHO) guidelines [24], and COVID-19 deaths, also assessed per WHO guidelines [25].

Every PCR test conducted in Qatar, regardless of location (outpatient clinic, drive-thru, or hospital, etc.), is classified on the basis of symptoms and the reason for testing (clinical symptoms, contact tracing, random testing campaigns, individual requests, healthcare routine testing, pre-travel, and at port of entry). PCR-confirmed infections are classified as “symptomatic” if testing was done because of clinical suspicion due to symptoms compatible with a respiratory tract infection.

Classification of infections by variant type was informed by weekly rounds of viral genome sequencing and multiplex, real-time reverse-transcription PCR (RT-qPCR) variant screening [26] of randomly collected clinical samples [15-19], as well as by results of deep sequencing of wastewater samples [17]. Based on existing evidence [27-29] and confirmation with viral genome sequencing [21], a B.1.1.7 case was defined as an S-gene “target failure” using the TaqPath COVID-19 Combo Kits (Thermo Fisher Scientific, USA) [30]. This accounted for >85% of PCR testing in Qatar, applying the criterion of a PCR cycle threshold (Ct) value ≤30 for both the N and ORF1ab genes, and a negative outcome for the S gene [29]. This definition was used to derive the B.1.1.7 case series data that were used subsequently to derive for only the B.1.1.7 variant.

### Empirical estimation methods

Five methods [13,31] were used to calculate 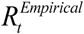 from daily diagnosed cases. To minimize effects of bias due to variation in the PCR testing volume over time, 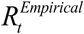 was calculated using only the time series of cases diagnosed due to presence of clinical symptoms. Cases diagnosed through testing conducted for other reasons were not used in these analyses, except in a sensitivity analysis.

#### Robert Koch Institute method

This method, which was chosen as the standard method for 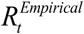 estimation in Qatar, utilizes the generation time (*τ*_*G*_), the time interval between the infection of an infector and an infectee in a transmission pair [13,22], to provide an estimate for 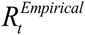. 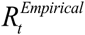. is calculated as the sum of newly diagnosed cases during *τ*_*G*_ consecutive days over the sum of previously diagnosed cases during the *τ*_*G*_ preceding days [22]. *τ*_*G*_ was assumed to be seven days, as informed by empirical evidence [32,33]. To smooth the curve and to avoid strong daily variations due to noise, 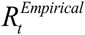 was calculated as a three-day moving average.

The range of uncertainty in the estimated 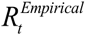 due to sampling variation was derived by applying the binomial sampling distribution to the number of positive PCR tests out of all tests, day by day, and repeating this process 1,000 times.

Four sensitivity analyses on the estimated 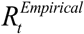 were conducted. In the first sensitivity analysis, the time series of all diagnosed cases (regardless of reason for PCR testing) was used instead of the time series of only symptomatic cases. In the second and third sensitivity analyses, the time series of hospital admissions in acute-care beds and ICU-care beds was used to proxy the epidemic trend, instead of the time series of symptomatic cases. In the fourth sensitivity analysis, the generation time *τ*_*G*_ was assumed to be 5, 7, and 10 days, instead of the fixed value of seven days [32,33].

#### Cislaghi method

This method utilizes the incubation time (*τ*_*I*_), the time interval between infection and symptom onset in an infected individual [33], to generate an estimate for 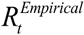. 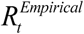 is calculated as the number of newly diagnosed cases on day *s* over the number of newly diagnosed cases on day *s* − *τ*_*I*_ [34]. *τ*_*I*_ was assumed to be five days [32,33]. To smooth the curve and to avoid strong daily variations due to noise, 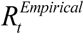 was calculated as a three-day moving average.

#### Wallinga and Teunis method

This method utilizes the serial interval (*τ*_*S*_), the time interval between symptom onset of an infector and that of an infectee [33], to generate an estimate for 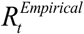. A likelihood-based estimate for 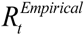 is derived by using pairs of diagnosed cases and the probability distribution for *τ*_*S*_ [35]. *τ*_*S*_ was assumed to have a Weibull distribution with a mean of 5.19 days and a standard deviation of 1.39 days, as informed by a meta-analysis of available data for SARS-CoV-2 infection [36].

#### Systrom-Bettencourt and Ribeiro method

This method utilizes an approximate relationship between 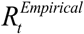 and the exponential growth of the epidemic, and assumes that 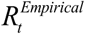 evolves as a Gausian process to provide a Bayesian 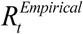 estimation [12,37-39]. A Gaussian filter was applied to account for daily variations (noise) in 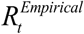 using a variance that was estimated by maximizing the log-likelihood of observing newly diagnosed cases [12,37-39].

#### Cori et al. method

This method utilizes the infectivity profile (*ω*_*S*_) of an infected individual to generate an estimate for 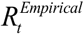 [31]. The average 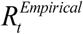 is estimated by the ratio of the number of newly diagnosed cases at time step *t*, to the sum of newly diagnosed cases up to time step *t* − 1, weighted by *ω*_*S*_. The infectivity profile was approximated by the distribution of the serial interval [31]. Bayesian statistical inference based on a Poisson process was used to generate the posterior distribution of 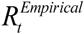, after assuming a gamma prior distribution for 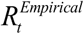 [31].

### Model-based estimation method

An age-structured deterministic mathematical model was developed to describe SARS-CoV-2 transmission dynamics in the population of Qatar [6,23]. The model was developed as informed by other models [6,23,40-42], and has been used, expanded, and continuously refined since the onset of the epidemic. This model has been the reference model for policy decision-making in Qatar, for providing forecasts, investigating epidemiology, and assessing the impact of public health interventions [6,23].

The model stratified the population into compartments according to age group (0-9, 10-19, 20-29, …, ≥80 years), infection status (infected, uninfected), infection type (asymptomatic/mild, severe, and critical), COVID-19 disease type (severe or critical disease), and vaccination status (vaccinated, unvaccinated) using sets of coupled, nonlinear differential equations (Figure S1 in the Online Supplementary Document).

The model was parameterized using current data for SARS-CoV-2 natural history and epidemiology [6,23]. It was fitted to the national standardized, integrated, and centralized databases of SARS-CoV-2 diagnosed cases, SARS-CoV-2 PCR and antibody testing, COVID-19 hospitalizations, and COVID-19 mortality [6], as well as to data of a series of SARS-CoV-2 epidemiological studies in Qatar [5,21,43-48]. The size and demographic structure of the population of Qatar were based on data from Qatar’s Planning and Statistics Authority [5,14,49]. 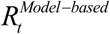 was derived using this model and was expressed in terms of the social contact rate in the population, transmission probability of the infection per contact, duration of infectiousness, and proportion of the population that is still susceptible to the infection [6,23]. A detailed description of the model, its input data, and fitting are available in References [6,23]. The model was coded, fitted, and analyzed using MATLAB R2019a [50].

### Agreements between *R*_*t*_ estimates

Agreements between different *R*_*t*_ estimates were assessed by calculating both the Pearson correlation coefficient, to assess the existence of a linear relationship, and also by calculating the Spearman correlation coefficient, to assess the existence of a monotonic (rank) relationship.

### Ethical approvals

The study was approved by the Hamad Medical Corporation and Weill Cornell Medicine-Qatar Institutional Review Boards with waiver of informed consent.

## RESULTS

The 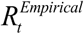 calculated using the Robert Koch Institute method captured effectively the evolution of the epidemic through its three waves, starting from the first wave (the wild-type variant wave) [5,6], the second (B.1.1.7) wave [15-19,21], and the third (B.1.135) wave [15-19,21] (Figure 1A). It also captured and correlated with key response landmarks, such as partial lockdowns during the three waves and subsequent easing of public health restrictions, and major social events that led to transient increases in the social contact rate in the population. It further captured transient fluctuations that were associated with significant clusters of infection, especially during low-incidence phases between August 1, 2020 and January 17, 2021, and between May 25, 2021 and August 18, 2021.

**Figure 1.**
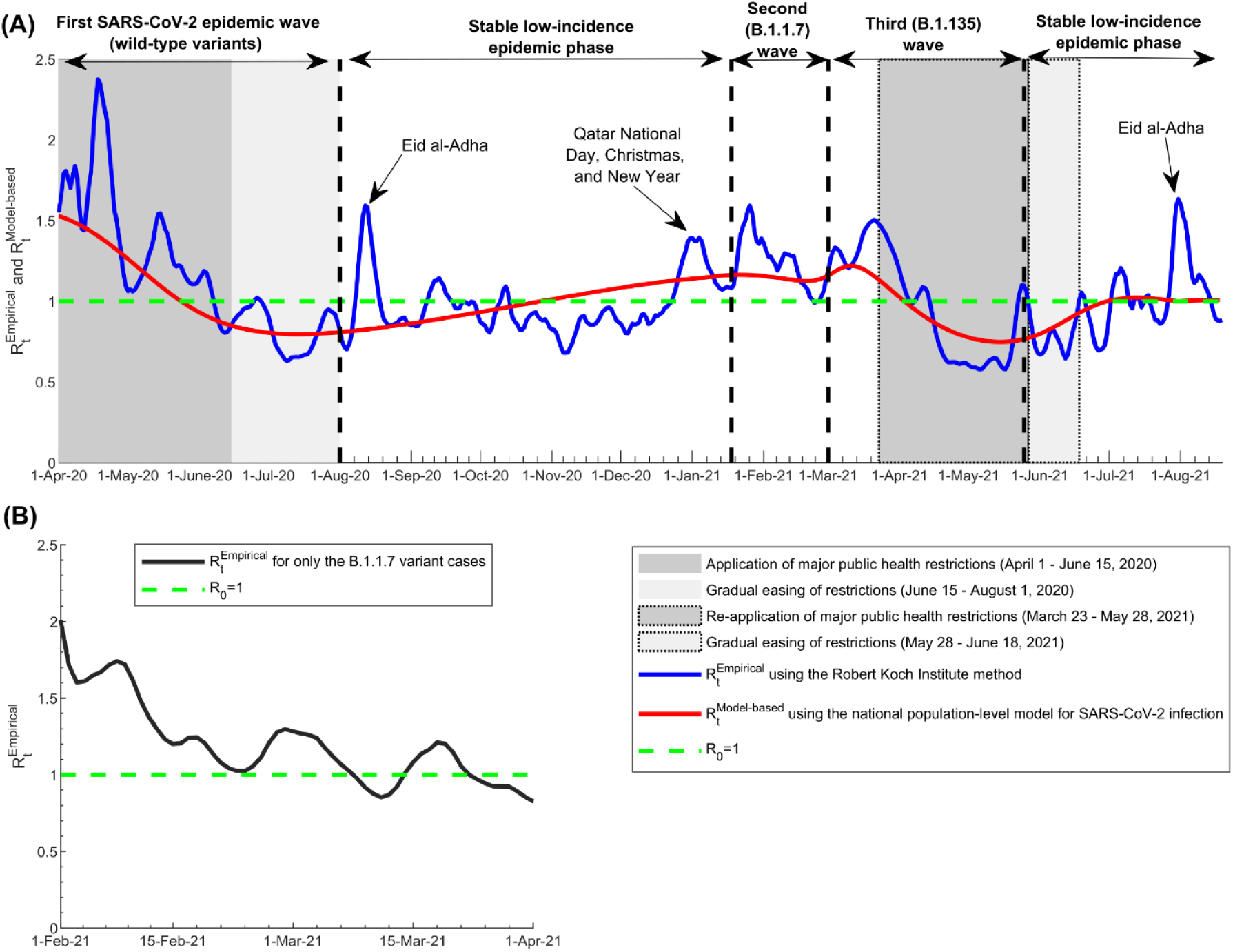
Effective reproduction numbers 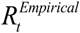 and 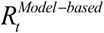 in Qatar. A) Trend in 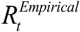 and 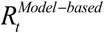, April 1, 2020 to August 18, 2021, and association with major events, response landmarks, and introduction and expansion of the B.1.1.7 and B.1.135 variants. B) Trend in 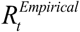 for only the B.1.1.7 variant cases, February 1, 2021 to April 1, 2021. 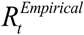 was estimated using the Robert Koch Institute method [22] applied to symptomatic case series data. The dashed green line represents the threshold of *R*_0_ = 1.

The epidemic expansion of B.1.1.7 cases starting from January 18, 2021 was associated with a large and rapid increase in 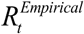 (Figure 1A), suggesting the higher infectiousness of this variant. 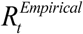 calculated using only B.1.1.7 case series data is shown in Figure 1B and demonstrated higher values, confirming further the higher infectiousness of this variant. 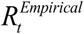 for only the B.1.1.7 variant averaged 1.45 during the exponential growth phase of the second (B.1.1.7) wave (February 1-22, 2021). It was unstable during the first two weeks of this wave (January 18-31, 2021; not shown), as transmission appears to have been influenced by one or more superspreading events that were not representative of the average community transmission. It was also unstable after April 1, 2021, as the number of daily B.1.1.7 cases was too small.

The first sensitivity analysis on the estimated 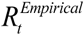, in which the time series of all diagnosed cases was used instead of only symptomatic cases, showed overall excellent agreement regardless of the input-data source used to calculate 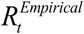 (Figure 2A). The Pearson correlation coefficient was 0.914 (p-value<0.001) and the Spearman correlation coefficient was 0.913 (p-value<0.001), both confirming the excellent agreement. There were only few noticeable differences that occurred when the number of diagnosed cases was too small (periods when SARS-CoV-2 incidence was low); thus, 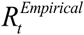 was more susceptible to transient variation in the number of diagnosed cases, such as due to sporadic, random PCR testing campaigns. Peaks in 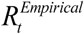 were also slightly larger using only symptomatic cases versus all diagnosed cases.

**Figure 2:**
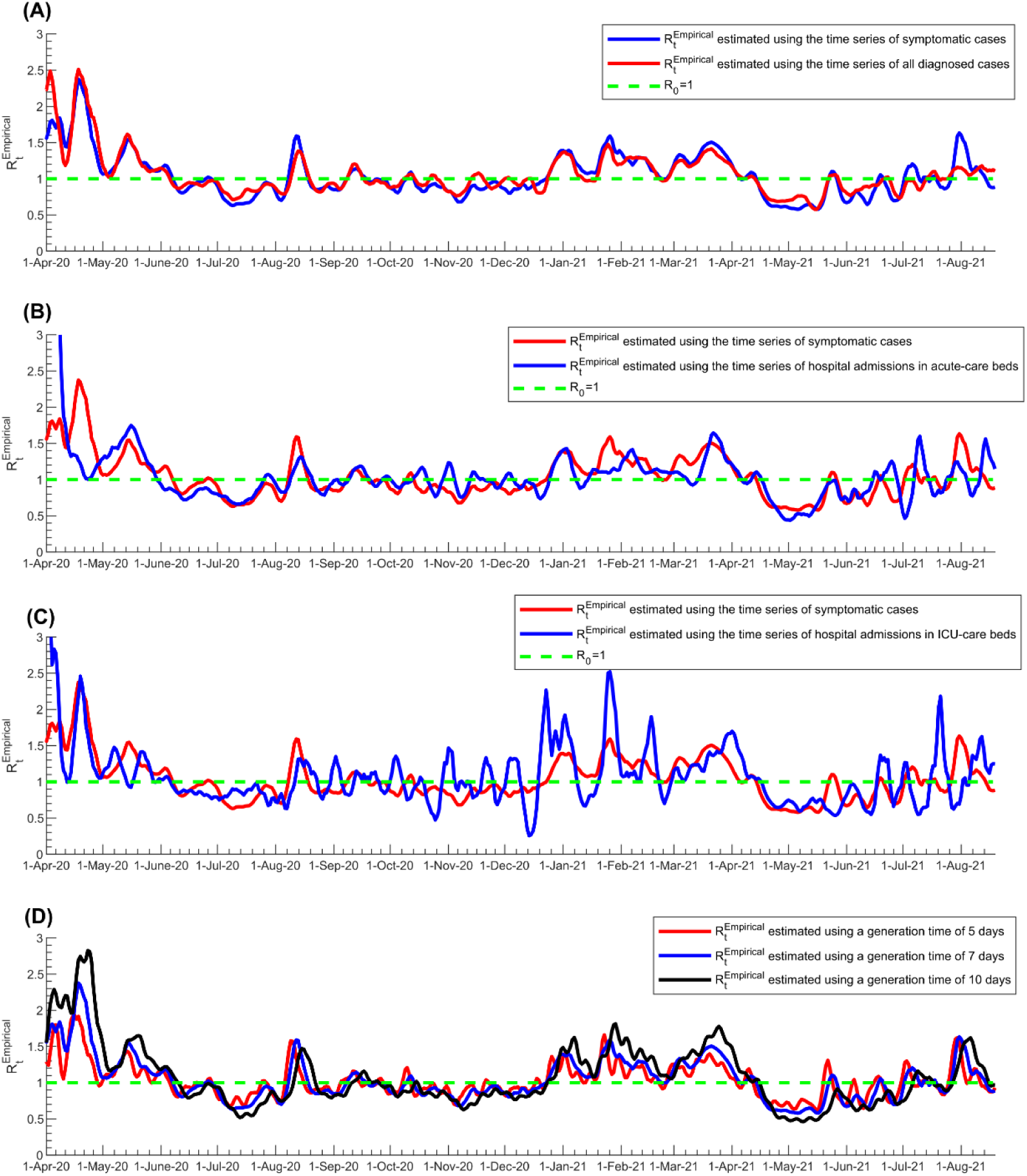
Sensitivity analyses of estimated 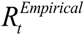 using the Robert Koch Institute method. A) Sensitivity analysis using the time series of all diagnosed cases instead of only symptomatic cases in estimating 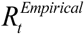. B) Sensitivity analysis using the time series of hospital admissions in acute-care beds instead of symptomatic cases in estimating 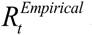. C) Sensitivity analysis using the time series of hospital admissions in ICU-care beds instead of symptomatic cases in estimating 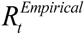. D) Sensitivity analysis using different values for the generation time in estimating 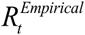. The dashed green line represents the threshold of *R*_0_ = 1.

The second sensitivity analysis, in which the time series of acute-care hospital admissions was used to proxy the epidemic trend, instead of the time series of symptomatic cases, showed rather strong correlation between 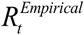 estimates (Figure 2B). The Pearson correlation coefficient was 0.512 (p-value<0.001) and the Spearman correlation coefficient was 0.716 (p-value<0.001), both confirming strong agreement. The third sensitivity analysis, in which the time series of ICU-care hospital admissions was used to proxy the epidemic trend, instead of the time series of symptomatic cases, also showed rather strong correlation between the 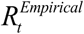 estimates, but also large fluctuations in 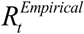 (Figure 2C). The Pearson correlation coefficient was 0.589 (p-value<0.001) and the Spearman correlation coefficient was 0.550 (p-value<0.001), both confirming strong agreement, but inferior to that for acute-care hospital admissions (Figure 2B versus Figure 2C).

The fourth sensitivity analysis, in which different values for the generation time *τ*_*G*_ were used, showed also strong agreement between different 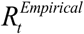 estimates (Figure 2D). The Pearson correlation coefficient was 0.901 (p-value<0.001) and the Spearman correlation coefficient was 0.900 (p-value<0.001), both confirming excellent agreement. The main differences between the estimates occurred in the timing and magnitude of peaks of the epidemic waves, as expected, since variation in generation time changes the rate of epidemic growth [51].

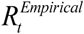 estimated using the Robert Koch Institute method (Figure 3A), Cislaghi method (Figure 3B), Systrom-Bettencourt and Ribeiro method (Figure 3C), Wallinga and Teunis method (Figure 3D), and Cori et al. method (Figure 3E), all showed similar results and were able to capture the evolution of epidemic waves and transient variations due to national public-health response landmarks and major social events. There were also overall strong correlations between them (Table 1 and Figure 4). However, the Systrom-Bettencourt and Ribeiro method (Figure 3C) tended to provide something of an average 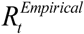 and was not as sensitive to transient changes in 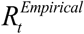 (Figures 3 and 4).

**Figure 3:**
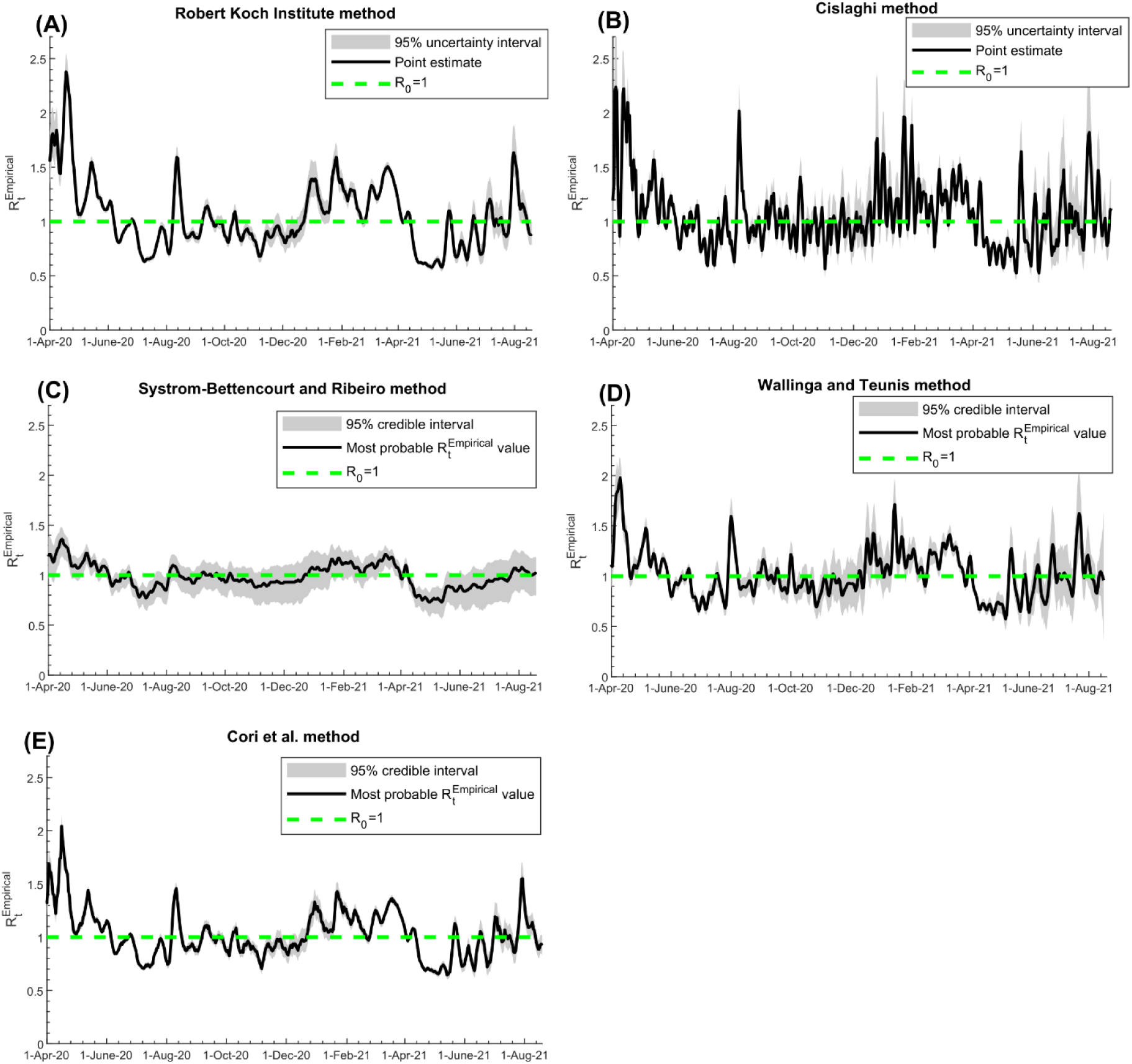
Trend in in 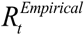 Qatar, April 1, 2020 to August 18, 2021, using the A) Robert Koch Institute method [22], B) Cislaghi method [34], C) Systrom-Bettencourt and Ribeiro method [12,37-39], D) Wallinga and Teunis method [35], and E) Cori et al. method [31]. The figure includes the 95% uncertainty or credible interval, as applicable for each method. The dashed green line represents the threshold of *R*_0_ = 1.

**Table 1.**
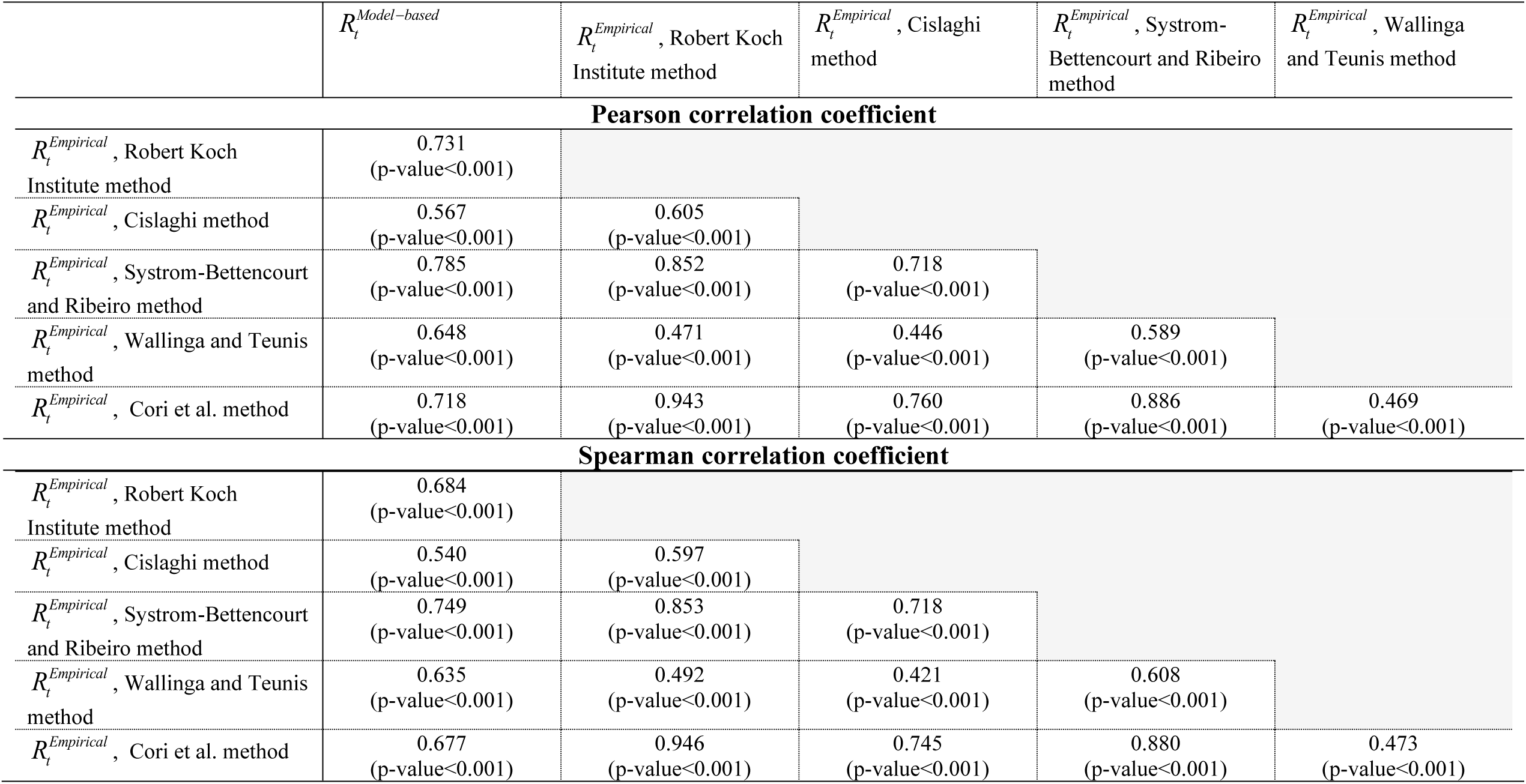
Correlations between 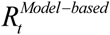 and 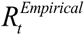 using the A) Robert Koch Institute method [22], B) Cislaghi method [34], C) Systrom-Bettencourt and Ribeiro method [12,37-39], D) Wallinga and Teunis method [35], and E) Cori et al. method [31].

**Figure 4:**
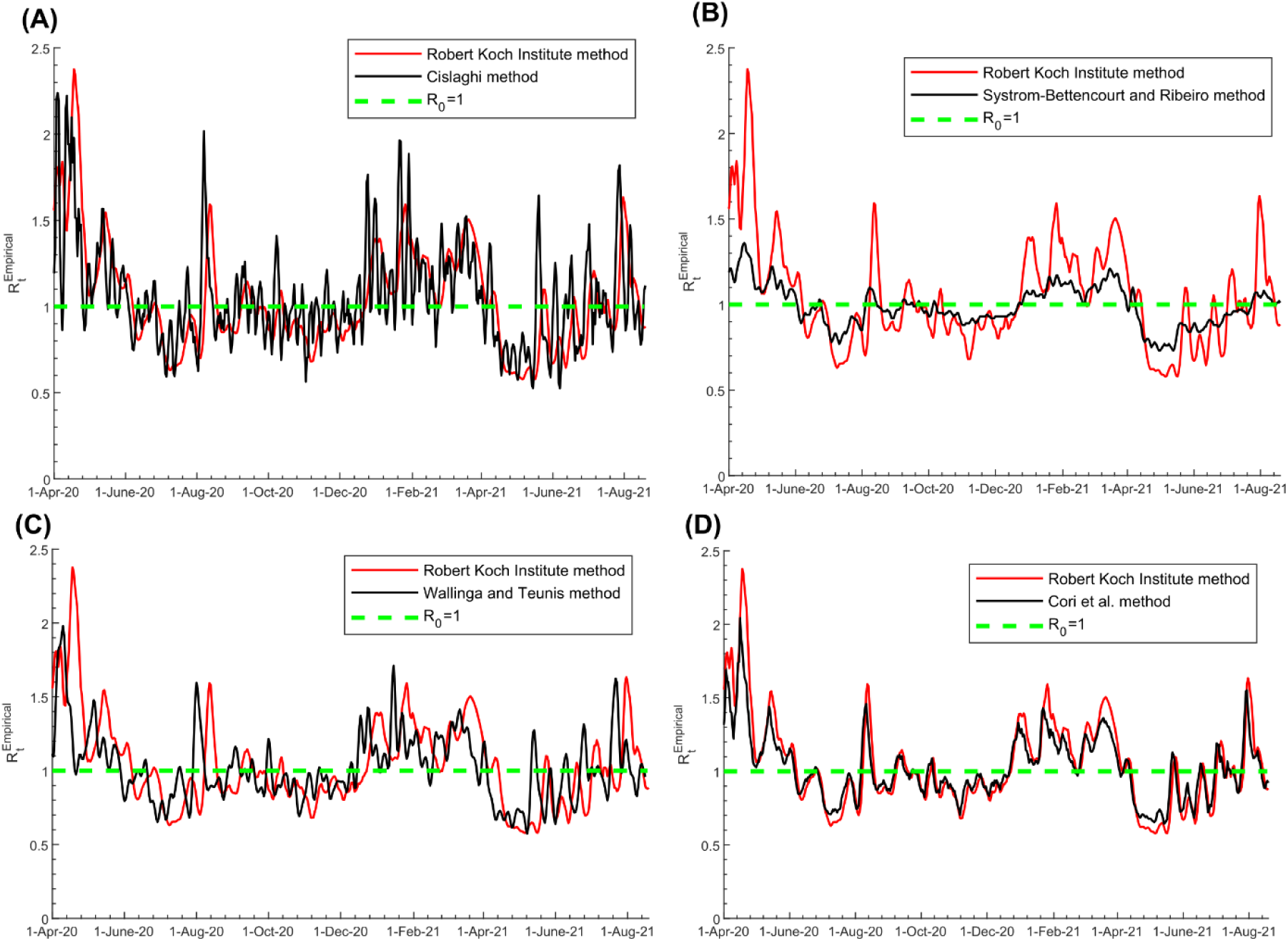
Pairwise comparison between 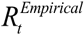 estimated using the Robert Koch Institute method [22] and that estimated using the A) Cislaghi method [34], B) Systrom-Bettencourt and Ribeiro method [12,37-39], C) Wallinga and Teunis method [35], and D) Cori et al. method [31]. The dashed green line represents the threshold of *R*_0_ = 1.

There were differences in how rapidly each method detected a change in epidemic dynamics (Figure 4). The Wallinga and Teunis method was the fastest at detecting a change, while the Robert Koch Institute method was the slowest, leading to weaker Pearson and Spearman correlation coefficients between them (Table 1). For instance, the surge in 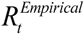 during the first Eid al-Adha after epidemic onset (a festival that occurred between July 30 and August 6, 2020 and is associated with celebrations and social gatherings) was detected on August 1, August 7, August 8, August 11, and August 13 using the Wallinga and Teunis, Cislaghi, Systrom-Bettencourt and Ribeiro, Cori et al., and Robert Koch Institute methods, respectively.

Uncertainty intervals around the 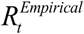 estimates of the various methods were narrow overall, except when the number of diagnosed symptomatic cases or the number of PCR tests was small, specifically during the low-incidence phases of the epidemic (Figure 3). Overall, the uncertainty in 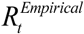 estimates did not impact the interpretation of the 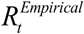 results (Figure 3). The only exception was for the Systrom-Bettencourt and Ribeiro method, as it showed rather wide uncertainty intervals compared to the point estimates for 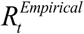 (Figure 3C).

The 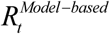 correlated strongly with the 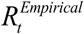 using different methods (Table 1), and provided somewhat of an average of the 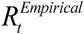 (Figure 1). For example, 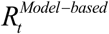 and 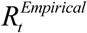 averaged 1.15 and 1.14 during the first wave, respectively. While it captured the three epidemic waves, it could not capture the transient fluctuations in 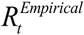, nor the effects of significant clusters during low-incidence phases.

## DISCUSSION

Two forms of *R*_*t*_ estimation were used in Qatar to inform the national COVID-19 response, the “empirical” estimation, 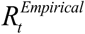, and the model-based estimation, 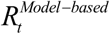. Both forms proved useful in real-time tracking of epidemic trends, understanding epidemic dynamics, and setting interventions to control transmission, such as application or easing of public health restrictions. Both forms were integral to the national public health response and to formulating evidence-based policy decisions to minimize the epidemic’s toll on health, society, and the economy throughout the phases of this epidemic.

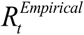 effectively captured the evolution of the epidemic during its three waves, the effects of the response landmarks, such as the partial lockdowns and easing of public health restrictions, and the major social events that affected the social contact rate in the population. Even transient fluctuations in infection transmission that occurred because of significant infection clusters were captured by 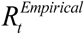. Strikingly, the introduction and expansion of the B.1.1.7 variant [21], that resulted in the second epidemic wave, was discovered immediately through 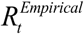 monitoring, as there was a sudden large, sustained increase in *R*_*t*_ that coincided precisely with a rapidly increasing number of S-gene “target failures” in PCR testing, even before viral genome sequencing was conducted to confirm the presence and expansion of this variant in the population [18].

While 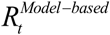 provided an average *R*_*t*_ that closely tracked the average 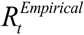, it did not have the resolution to capture transient changes in 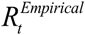, other than major changes associated with the three epidemic waves. Still, 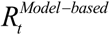 was useful and influential, as it was, along with the model that generated it [6,23], the basis for forecasting and future planning, such as forecasting the epidemic time-course and epidemic potential, forecasting healthcare needs of acute-care and ICU-care bed hospitalizations, predicting the impact of social and physical distancing restrictions, planning for easing of restrictions, and forecasting the impact of different mass vaccination strategies [6,23]. Therefore, both forms of *R*_*t*_ complement each other and should be part of any effective COVID-19 national response.

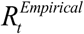 estimation proved robust in sensitivity analyses conducted to assess its utility. Baseline estimation of 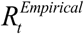 was based on the time series of symptomatic cases as a proxy of the actual incidence of SARS-CoV-2 infection in the population, which is unknown. Using the time series of all diagnosed cases instead of just symptomatic cases did not appreciably impact 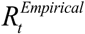 estimation, even though PCR testing volume and strategies varied throughout the epidemic. Using the time series of acute-care hospital admissions instead of the time series of symptomatic cases also led to comparable estimates for 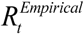. This was also the case, but with weaker agreement, when the time series of ICU-care hospital admissions was used to proxy trends in infection incidence. This is not surprising as there is a long delay between onset of infection and ICU-care hospital admission, and the number of ICU-care admissions was relatively small with the low COVID-19 severity in Qatar’s predominantly young and working-age population [5,48]. Variations in the 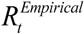 assumed value for the generation time in the estimation did not heavily impact estimates. These findings support the robustness of the approach employed to estimate 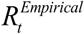.

Examination of different methods to estimate 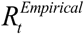 demonstrated consistency of the results, generally strong correlations between the estimates, and an acceptable level of uncertainty in them. The only exception was the Systrom-Bettencourt and Ribeiro method which tended to provide something of an average 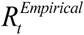. It was not as sensitive to transient changes in 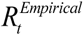, and had wide uncertainty intervals compared to the point estimates. There were also differences in how rapidly each method detected a change in epidemic dynamics. The Wallinga and Teunis method was the fastest to detect a change, while the Robert Koch Institute method was the slowest. Yet, overall, these findings support the robustness of using these methods in 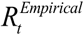 estimation and to guide COVID-19 national responses.

This study has limitations. The estimated 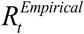 and 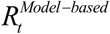 are contingent on the validity and generalizability of input data. The uncertainty/credible intervals estimated here accounted for the uncertainty arising from sampling variation, or from our imperfect knowledge of specific epidemiological quantities, such as the serial interval, but did not account for other sources of uncertainty, such as our imperfect knowledge of the true incidence of infection in the population. To reduce bias due to variation in volume and strategies of PCR testing over time, 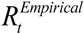 was calculated using the time series of symptomatic cases, but the distribution of the delay between onset of infection and onset of symptoms may bias these estimates. 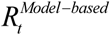 was estimated using a deterministic compartmental model, but this type of model may not be representative of stochastic transmission dynamics, particularly when the number of infections is small. Despite these limitations, 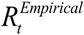 and 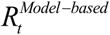 were able to capture the evolution of the epidemic through its several waves, and to effectively inform the national response and policy decision-making.

## CONCLUSIONS

*R*_*t*_ estimations played a critical and influential role in the COVID-19 national response in Qatar. Even though surveillance data of SARS-CoV-2 infection are imperfect and prone to bias, *R*_*t*_ estimations were robust and generated consistent results regardless of the data source used, or the method employed in generating estimates. These findings affirm the value and complementarity of using both 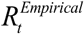 and 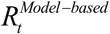 to track the epidemic in real-time and to inform public health decision making at a national level. This can also be done despite low-resource demands, as *R*_*t*_ estimation utilizes existing surveillance data. Moreover, application of some of the estimation methods is feasible even without established expertise in infectious disease modeling. Since the choice of estimation method does not impact the estimates, each country may decide on the best approach, method, and source of data to be used in the estimation, weighing feasibility, ease of use, and functionality, given its specific circumstances.

## Data Availability

The dataset of this study is a property of the Qatar Ministry of Public Health that was provided to the researchers through a restricted-access agreement that prevents sharing the dataset with a third party or publicly. Future access to this dataset can be considered through a direct application for data access to Her Excellency the Minister of Public Health (https://www.moph.gov.qa/english/Pages/default.aspx).

## Acknowledgements

HHA and RB acknowledge the joint support of Qatar University and Marubeni M-QJRC-2020-5. The authors are grateful for support provided by the Biomedical Research Program and the Biostatistics, Epidemiology, and Biomathematics Research Core, both at Weill Cornell Medicine-Qatar, as well as for support provided by the Ministry of Public Health and Hamad Medical Corporation. The developed mathematical models were made possible by NPRP grant number 9-040-3-008 (Principal investigator: LJA) and NPRP grant number 12S-0216-190094 (Principal investigator: LJA) from the Qatar National Research Fund (a member of Qatar Foundation; https://www.qnrf.org). The authors are also grateful for the Qatar Genome Programme for institutional support for the reagents needed for the viral genome sequencing. The statements made herein are solely the responsibility of the authors. The funders had no role in study design, data collection and analysis, decision to publish, or preparation of the manuscript.

## Author contributions

RB and HHA constructed, coded, and parameterized the mathematical models, and conducted the analyses. HC conducted statistical analyses. HHA and LJA. conceived and led the design of the study, construct and parameterization of the mathematical models, and wrote the first draft of the article. All authors contributed to discussion and interpretation of the results and to the writing of the manuscript. All authors have read and approved the final manuscript.

## Supplementary Material

**Figure S1.**
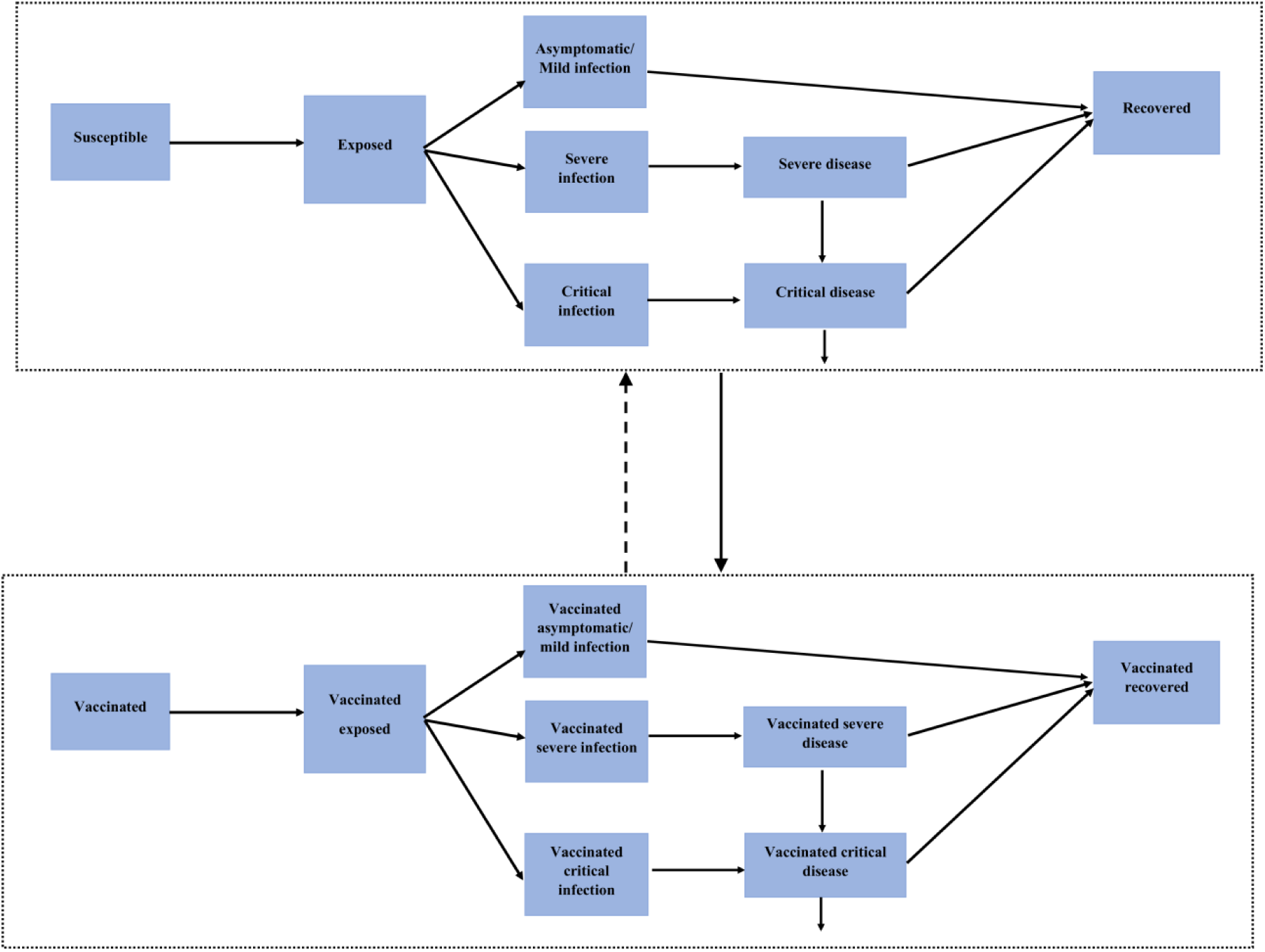
A conceptual diagram illustrating the basic structure of the deterministic mathematical model developed to describe SARS-CoV-2 transmission dynamics in the population of Qatar. The detailed structure of this model and its description are found in [1,2]. In this figure, solid lines denote progression or forward movement from one population compartment to the next, while dashed lines denote backward movement from the present population compartment to the previous one.

